# Epidemiology and medical costs of orthopedic conditions in a tertiary hospital in Kenya; A five-year analysis of admission data

**DOI:** 10.1101/2022.03.20.22272413

**Authors:** Elesban Kihuba

**Author notes:** Corresponding author Elesban Kihuba.

## Abstract

**Background:** Orthopedic services are a key component of the surgical healthcare system. Scaling up these services would not only reduce the musculoskeletal diseases burden but also the overall disease burden significantly. As such quality intelligence on epidemiology and costs of orthopedic services should be accurately represented for guiding effective policy formulation and implementation.

**Aim:** The aim of the study was to provide a snapshot of the pattern of orthopedic admissions, care and associated medical costs in Kenya.

**Methods:** A retrospective study was conducted reviewing case files of patients admitted to the orthopedic department from 1^st^ January 2015 to 30^st^ November 2019.

**Results:** There were 4320 distinct musculoskeletal admission diagnoses among the 3527 admissions. The study shows that trauma accounts for 53.04% of all admissions while joint replacement, shoulder and knee, limb deformities, infection, removal of hardware, ankle and foot and tumour procedures accounts for 15.01%, 8.87%, 7.29%, 6.44%, 3.94% and 3.23% respectively. Joint replacement is the leading diagnosis among patients in the 61 to 75 years age group and fragility fractures and posttraumatic arthritis featured prominently as the key reasons for joint reconstruction. The proportion of admissions with health insurance (60.82%) was exceptionally high compared to the national average (19.59%). However, current financing mechanism provide inadequate social protection against cost of orthopedic surgery.

**Conclusion:** The case volume and mix highlight the growing importance of trauma, fragility fractures and degenerative joint diseases in orthopedic care in developing country set up. Overall, the observed model of care and associated outcome and costs provide a blueprint for orthopedic system development.

## 1. Introduction

Provision of quality and affordable surgical services is now an instrumental goal for the healthcare system^1,2^. The milestone path was checkered with new and mounting evidence that surgical burden account for close to 30% of the global burden of diseases and provision of basic surgical interventions can avert close to 3.2% and 5.5% of annual mortality and morbidity respectively^34^.

Orthopedic and trauma conditions contribute significantly to the surgical burden and comprise the second highest global volume of years lived with disability^5^. Further, musculoskeletal burden increased by 61.6% from 1990 to 2016^5,6^. This reinforces the need to strengthen orthopedic and trauma care as part and parcel of essential packages for universal health coverage^7–10^. Indeed, interventions to address orthopedic and trauma are cost effective: median cost effectiveness ratio (CER) for orthopedic surgery (USD 381.15 per DALY) is excellent compared with a number of chronic medical conditions including ischemic heart disease and HIV treatment^4^.

But while data indicates that low and middle income countries are disproportionately affected by musculoskeletal and trauma conditions, these countries lag behind in their response towards strengthening orthopedic and trauma care^7,9^. With the prominent challenge being “how to expand health services to meet growing needs with limited resources’^11,12^. As such quality data is a key ingredient in providing ‘local answers’ and a road map to guide the orthopedic surgical system development.

In Kenya, research activity on orthopedic and trauma services is negligible^13^. Further, little is known of the epidemiology and cost of procedures for orthopedic and trauma conditions^14^. This article is an attempt to bridge this gap in data by examining characteristics of orthopedic admissions, a description of care provided and associated direct medical costs of all admissions over a five-year period (2015-2019), to a level five health facility.

## 2. Methods

### 2.1 Setting

This was a retrospective study conducted at an orthopedic and rehabilitation department of a leading Mission hospital in Kenya. It is a 33-bed capacity department, and the patients are under the care of seven (7) orthopedic specialists, three (3) orthopedic residents, a physician assistant and twenty (20) nurses. There are two operating rooms at the department, and they are operational on weekdays between 9AM and 5 PM. These are equipped with basic facilities for major surgery but lack theatre ventilation systems. The orthopedic department also provides outpatient services on weekdays and attends to an average of 18,737 clients per year.

### 2.2 Data collection and analysis

Data collection was conducted by a team of 5 research assistants over a six-month period from July 2019 to December 2019. The team underwent training including conducting a pilot study. All patients admitted for surgery from January 2015 to November 2019, irrespective of their diagnosis, were included in the study. Data abstraction was done by reviewing each patient file and directly transcribed into an online interface based on KoBotoolbox (an open-source platform for form design and data collection). Data collected from the paper-based records did not contain any identifiable information which would allow attribution of private information to an individual.

At time of entry, data were subjected to skip and validation to minimize errors. In addition, the authors conducted periodic audits. Data collected falls under the following categories: sociodemographic; admission notes through the patient pathway (reason for admission, mechanism of injury, duration of symptoms, time to admission, comorbidity, investigations); clinical diagnosis; perioperative care; surgical intervention; discharge outcome; and direct medical costs (out of pocket payment and insurance reimbursement). For standardization purposes, admission diagnosis was sub-classified by orthopedic subspecialties: Trauma; Ankle and foot; Spine; Arthroplasty; Infections and neoplasm. At the end of the data collection exercise, the data set was then exported to STATA for analysis.

Categorical variables are reported as a proportion or percentage of the study population while continuous variables are reported as median and interquartile range (IQR) stratified by age and specialties.

The cost lists for orthopedic procedures were estimated from the hospital perspective. The cost list includes the following economic information: the average cost per diagnosis category, the average length of stay and total number of cases. The average costs under this study are defined to include all the costs generated during hospitalization: admission process costs, admission days, surgical procedures costs, laboratory tests and imaging costs. Both the out of pockets payments and insurance contributions were included in the cost analysis.

### 2.3 Ethics

The research protocol for this study was reviewed and approved by the Hospital research and ethic committee. The Hospital administration also approved the study including its publication.

## 3. Results

### 3.1 Basic characteristics

Socioeconomic characteristics of all admissions are provided in Table 1. Data for 3527 patients were analyzed, of which 59.35% were male while 40.65% were female. The median age (IQR) at admission was 40 (25-59). The data shows that the productive age groups and the elderly are disproportionately affected. The pediatric cases accounted for 18.84% of all cases, while the 19-24, 25-44, 45-60, 61-79 and over 80 years age groups accounted for 5.81%, 32.90%, 19.95%, 18.33% and 4.11% respectively. The proportion of patients with health insurance (60.82%) was exceptionally high compared to the national average (19.59%), with 96% of patients with insurance cover subscribing to the national health insurance fund (NHIF) compared to 4% with private health insurance covers. The unemployment level was lower than the national average.

**Table 1:** Basic characteristics

| Basic characteristics | Study Data | National |
| --- | --- | --- |
| Sex n=3527 |  |  |
| Male n (%) | 2,093<br>(59.35%) | 49.51% |
| Female n (%) | 1,434<br>(40.65%) | 50.49% |
| Age n=3527 |  |  |
| Median (IQR) | 40 (25-59) | 20 |
| 0-18 | 665 (18.84%) | 48.05% |
| 19-24 | 205 (5.81%) | 11.39% |
| 25-44 | 1161 (32.90%) | 25.93% |
| 45-60 | 704 (19.95%) | 9.42% |
| 61-79 | 647 (18.33%) | 4.33% |
| >80 | 145 (4.11%) | 0.872% |
| Occupation |  |  |
| Unemployed | 721(20.42%) | - |
| Self-employed | 645 (18.27%) | - |
| Agriculture | 496 (14.05%) | - |
| Retired | 197 (5.66%) | - |
| Civil servants | 111 (3.44%) | - |
| Paed/student | 706 (20.02%) | - |
| Others | 651(18.48) | - |
| Insurance |  |  |
| Health insurance | 2145(60.82%) | 19.59% |
| None | 1382(39.18%) | - |

### 3.2 Epidemiology of orthopedic admission

There were 4320 distinct musculoskeletal admission diagnoses among the 3527 admissions (2914 new patients and 613 readmissions). In total, 4204 orthopedic operations were performed. An analysis of a patient’s admission diagnosis by orthopedic subspecialties stratified by age is represented by Table 2.

**Table 2:** Distribution of admissions by orthopedic specialties and age groups

| Diagnosis | Overall | <18 | 19-24 | 25-44 | 45-60 | 61-79 | >80 |
| --- | --- | --- | --- | --- | --- | --- | --- |
| Comorbidity rate | 20.18% | 2.10% | 1.48% | 9.09% | 26.42% | 49% | 49.66% |
| Trauma | 53.04% | 46.24% | 62.44% | 64.63% | 54.40% | 34.31% | 62.70% |
| Total hip replacement (THR) | 9.70% | - | - | 2.41% | 10.37% | 33.69% | 20.69% |
| Elbow, Shoulder, Knee | 8.87% | 11.11% | 17.56% | 13.25% | 11.22% | 3.55% | 1.38% |
| Limb deformities | 7.29% | 28.08% | 6.83% | 3.27% | 3.41% | 1.24% | - |
| Infections | 6.44% | 7.51% | 7.80% | 8.9% | 6.82% | 5.1% | 4.14% |
| Total knee replacement (TKR) | 5.31% | - | - | 0.69% | 6.39% | 18.39% | 9.65% |
| Removal of hardware (ROH) | 3.94% | 5.67% | 7.35% | 6.99% | 4.60% | 3.44% | 2.78% |
| Ankle and foot | 3.23% | 1.65% | 2.44% | 5.42% | 5.54% | 2.16% | - |
| Tumors | 1.64% | N/A |  |  |  |  |  |
| Spine | 0.6% | N/A |  |  |  |  |  |

Trauma is the leading cause of admission across all age groups. Trauma accounts for 53.04% of all admissions while joint replacement, elbow/shoulder/knee, limb deformities, infection, ROH, ankle/foot and tumour procedures accounts for 15.01%, 8.87%, 7.29%, 6.44%, 3.94% and 3.23% respectively. The first spine surgery case was performed in mid-2019.

Among the trauma patients, data shows higher trauma rates for male (60.63%) than female; for right sided (54.42%) than left sided extremities; for patients 19-44 years old and over 80 years old. The median age for trauma patients was 38 (25-53). The proportion of trauma cases with an open injury was 12.49% while fragility type fractures accounted for 82% and 50.45% of the fractures in the over 80 years and 61-79 years group. Only 7.22% of the patients were treated within 48 hours after the injury. Indeed 39.29% of all injuries were treated after 21 days of injury. Road traffic accidents, falls and fall from a height were the leading mechanism of injury.

Among the older age groups (>61 years), THR and TKR were among the most common admission diagnoses. The median (IQR) age of the patient undergoing total hip replacement and total knee replacement was 67 (58-73) and 65 (59-71) respectively. Most patients who had hip and knee replacement (65.15% and 80.21% respectively) were female. THR causes were distributed as follows: primary arthritis (53.30%), neck of femur fracture (19.58%), posttraumatic arthritis (18.99%), failure of prosthesis (5.93%), avascular necrosis (1.19%) and infection (0.88%). In the TKR group, inflammatory arthritis, primary arthritis, posttraumatic arthritis, failure of prosthesis and infection accounted for 46.28%, 35.64%, 12.77%, 3.19% and 2.13% of all causes, respectively. Of all TKR patients, 12.50% had fixed flexion deformities, 62.50% and 25% had valgus and varus deformities.

Removal of hardware and infections accounts for 10% of the admissions. On infections, fracture related infection (38.93%), chronic osteomyelitis (35.25%), hip septic arthritis (6.56%), knee septic arthritis (5.37%) and acute osteomyelitis (2.87%) featured among the most common types of infection.

### 3.3 Preoperative management

A preoperative checklist has been adopted to foster provision of preoperative care. Basic laboratory investigations conducted for all admissions-both adult and pediatric cases-included a full blood count and a urea, electrolytes, creatinine test and random blood sugar. Specific investigations include: HbA1c; group and cross matching for major surgeries; Echocardiogram and electrocardiography for male and female patients above 55 years and 60 years respectively or with known chronic illnesses; C-reactive protein (CRP) and erythrocyte sedimentation rate (ESR) for all suspected infected cases.

The rate of comorbidities was higher among female patients (30.89%) and increased with age: from a low of 2.10% in the age group under 18 and 1.48% in the group 19 to 24, rising to a high of 49.66% among patients older than 80 years. Data on obesity was unavailable and thus has been excluded from the analysis. HIV and asthma were the leading cause of comorbidities in the group younger than 44 years while diabetes mellitus, hypertension or combination of diabetes and hypertension were the leading coexisting comorbidities among patients older than 45 years. Of the 2273 patients who had a HIV test, only 4.89% (106) turned out positive. The preoperative assessment was two pronged; a preoperative assessment for all patients and then a thorough review by a medical specialist for those at risk. All the patients deemed unfit for surgery after the assessment were referred to other higher facilities for services.

### 3.4 Intraoperative management

On chemoprophylaxis, a single dose of ceftriaxone was the drug of choice. This was administered in the operating room. In the adult population group, choice of anesthetic technique was based on type of surgery. For example, all joint replacement and lower limb procedures were carried out under spinal anesthesia. On the other hand, most, pediatric cases were safely anaesthetized with general anesthetic technique (60.51%).

With regards to wound management-all surgical wounds with exception of the infected ones were closed in layers with the skin closed in two layers-a subdermal and subcuticular closure with 2-0 monofilament suture and a compression dressing applied.

### 3.5 Postoperative management

Prophylactic parenteral antibiotics- ceftriaxone- was administered twice a day till the day of discharge and then oral cefuroxime continued for at least 7 days. On wound care, all soiled dressing is changed at discharge and then opened at the second week postoperatively. The immediate postoperative mortality rate was low- (0.21% or 7 patients) and all were sudden unexpected deaths while 10% were lost to follow up at 6th week.

### 3.6 Cost data

The average costs of procedures are shown in Table 3. The proportion of orthopedic procedure costs covered by third parties differed widely by surgical diagnoses. Joint replacement enjoyed the best reimbursement rates while trauma, tumour, limb deformities and infection attracted a reimbursement rate lower than 50% of the average direct medical costs. All the admitted patients experienced prolonged hospital stay.

**Table 3:** Postoperative outcomes and costs by orthopedic specialties

| Orthopedic category | Median (IQR) duration of operation in minutes | Median (IQR) Length of stay | Average cost of procedure | Proportion reimbursed by Insurance |
| --- | --- | --- | --- | --- |
| Trauma | 90 (60-120) | 6(3-9) | 149273.7<br>(USD<br>1471.59) | 39.54 |
| THR | 120 (90-140) | 7(5-8) | 328991.8<br>(USD<br>3243.33) | 80.99 |
| Elbow,<br>shoulder,<br>Knee | 90 (60-120) | 4(2-6) | 137558.5<br>(USD<br>1356.10) | 66.02 |
| Limb<br>Deformities | 70 (60-120) | 5(3-8) | 113881.7<br>(USD<br>1122.69) | 42.52 |
| Infection | 60 (45-90) | 6(4-10) | 140231.6<br>(USD<br>1382.46) | 51.61 |
| TKR | 120 (90-120) | 5(4-7) | 386572.6<br>(USD<br>3811.00) | 62.43 |
| ROH | 60 (50-90) | 4(2-6) | 117663.4<br>(USD<br>1159.97) | 72.11 |
| Ankle &<br>foot | 90 (60-140) | 4(3-7) | 128027.1<br>(USD<br>1262.14) | 64.23 |
| Tumour | 47 (35-60) | 4(2-6) | 74102.35<br>(USD<br>730.53) | 36.75 |

USD Exchange rate at 101.436.

## 4. Discussion

Transforming orthopedic services in Kenya requires quality data on the burden and cost of orthopedic services. In this study, we examined the admission patterns, care and cost of orthopedic procedures among patients admitted with orthopedic and trauma conditions. The scope and variety of orthopedic conditions encountered, make a case for development of orthopedic healthcare delivery system including dealing with inefficiency issues and low funding for orthopedic care.

In terms of sociodemographic characteristics, our findings were consistent with other studies showing that the male group, the productive age groups and the elderly are disproportionately affected by musculoskeletal conditions^15,16^ with trauma being the leading cause of admission across all age groups. The study also indicated the following peculiarities of the characteristics of admissions: while the median age at admission was 40 years, the median age among the male group was lower-36 years than the female group-47years; right lower limb and left upper limb were predominantly affected; fragility like fractures rates are high in the over 61 years group; joint replacement is the leading cause of admission in the over 61 years group; and that one in five patients presented with a comorbidity.

The high rate of comorbidity among orthopedic patients over 61 years in our set up is worthy of attention and as such preoperative strategies should be instituted to optimize their wellbeing^17,18^. In the study hospital, optimization for the procedures was achieved through the two-pronged approach. On the other hand, while increasing elderly populations drive the burden of fragility fractures^19–21^, the higher rates observed in this study suggests a high burden of osteoporosis and is also of note due to non-existent preventative and treatment programs for osteoporosis in the country.

In regards to arthroplasty, the median age and gender ratios are comparable to that recorded in other studies^22,23^ but differs slightly from finding from a study from Malawi and Ghana^24,25^. The study however, points to an increased importance of fragility fractures or neck of femur fracture in total hip arthroplasty and posttraumatic arthritis in both total hip and knee arthroplasty.

The median length of stay observed in this study was longer compared to literature from the developed world^26^ but consistent with data from the developing world^22,24,27^. A possible explanation based on our experience could be due to little emphasis on cost effective care and resources miss match resulting in prolonged hospital stay and delay in accessing supporting services such as radiology, physiotherapy and laboratory.

On the model of care, procedure related costs were financed through health insurance and self-payment. Given the high out of pocket payments for orthopedic procedures, the findings call for cooperation between providers and insurers aimed at improving the efficiency of care through strengthened costing systems^29^ as envisaged in current policy ^30^. Further, the findings of access delay to definitive fixation for trauma related injuries, a Bellwether procedure, calls for improvement in access to orthopedic care in Kenya^31,32^. Further research is required to help us understand the extent and nature of barriers that limit access to orthopedic care.

While the hospital attends to over eighteen thousand clients a year, the analysis was limited to those cases that were severe to warrant surgical treatment. Thus, the characteristics and costs of cases treated conservatively were excluded. The retrospective nature of the study exposed it to weaknesses inherent in routine data sources. Further, the study was conducted in a well-established orthopedic hospital thus findings may not be generalizable to other facilities. It is our understanding that the data, however, sheds light to the different aspects of the orthopedic surgical health system that ought to be strengthened to improve quality and efficiency of care.

## 5 Conclusion

The study demonstrates the characteristics of admissions to an orthopedic and trauma hospital in Kenya. The case volume and mix highlight the growing importance of trauma, fragility fractures and degenerative joint diseases in orthopedic care in Kenya. On the other hand, insurance enrollment is high, but some common orthopedic procedures including trauma, infections and tumour related ones attract a high proportion of out-of-pocket payments. Two key sources of inefficiency noted in this study include the prolonged hospital stay and delayed treatment. Overall, the observed model of care and associated outcome and costs provide a blueprint for orthopedic system development. This information is especially vital given the ongoing discussions on universal health coverage in Kenya.

## Data Availability

All data produced in the present study are available upon reasonable request to the authors

## Acknowledgements

The authors are grateful to the Hospital management who gave permission for this work and provided resources for data collection. The authors thank the data collection team who worked tirelessly to abstract data into an electronic form. They are also grateful to the hospital’s health workers who daily provide care to orthopedic and trauma patients from all parts of Kenya.

## Funding

This work was supported by the Hospital and its partners who had no role in the research activities.

## Author contribution

All authors participated in the conceptualization, design and review of the article for publication. EK took lead in design of the methods, analysis and writing of the manuscript.

## Competing interest

The author declares they have no competing interests.

## Notes

### Competing Interest Statement

The authors have declared no competing interest.

### Funding Statement

The author(s) received no specific funding for this work. The research study was
nested in a quality improvement programme.

### Author Declarations

The research protocol for this study was reviewed and approved by PCEA Kikuyu Hospital research and ethic committee. The Hospital administration also approved the study including its publication.

